# Which factors influence decisions to withdraw from eculizumab: a qualitative study of patients diagnosed with aHUS

**DOI:** 10.1101/2025.07.21.25331504

**Authors:** Jan Lecouturier, Neil Sheerin

**Author notes:** **Corresponding author**: Professor NS Sheerin, National Renal Complement Therapeutic Centre, Translational and Clinical Research Institute, Newcastle University, Newcastle upon Tyne.

## Abstract

**Background:** Atypical haemolytic uremic syndrome (aHUS) is a rare life-threatening disease. Lifelong treatment with intravenous eculizumab every two/three weeks was recommended but evidence is emerging that patients can stop eculizumab and restart should they relapse. However, little is known about the opinions and needs of aHUS patients on withdrawal.

**Objective:** We aimed to understand the factors that impact on decisions to withdraw from treatment.

**Methods:** We analysed in-depth telephone interviews thematically using a constant comparative method. Interviewees included adults (8), and the parents of children (12), with aHUS approached to participate in an eculizumab withdrawal trial.

**Results:** The onset of aHUS had been traumatic for most. Regarding eculizumab, withdrawal group participants talked of the disruptive treatment regime and side effects, the time off work/school and impact on taking holidays. Decisions to withdraw from eculizumab were driven by the wish to lead more normal lives and concerns about long-term treatment. Drivers for declining withdrawal focused on relapse and its perceived impact. After two years the withdrawal group had regained a semblance of normality, though fears about relapse remained and they were aware of the need for long-term follow-up. Participants had a greater sense of control over the necessary steps should they/their child relapse.

**Conclusion:** Understanding patient/parent experiences should guide discussions about eculizumab withdrawal. Support to alleviate fears in the early stages of withdrawal would be beneficial. Evidence from the main trial on successful withdrawal, and recovery time where eculizumab was reinstated may provide reassurance to those who are uncertain about withdrawal.

**Key findings and implications:** Lifelong treatment can be burdensome for patients and families and have a substantial negative impact on their lives. This is particularly so when the onset of illness occurs, and treatment is commenced, in babies and young children.

Long-term treatment can be disruptive to work and school, due to the fortnightly administration of eculizumab and the side effects experienced. There was a dislike for long-term medication particularly when there was uncertainty as to whether treatment was still warranted. The opportunity to consider treatment withdrawal was welcomed.

Conversations between clinicians and patients/parents about withdrawal should be guided by an understanding of the experiences at onset and living on treatment, concerns and fears. Hearing parents’ and adults’ accounts of the positive impact of withdrawing from eculizumab may provide reassurance to others.

## 1 Introduction

Atypical haemolytic uremic syndrome (aHUS) is a rare life-threatening disease most frequently due to excessive activation of the complement system which leads to thrombus formation in small blood vessels, causing damage to internal organs, in particular the kidneys. The onset of aHUS is attributed to certain genetic, environmental, and immunologic factors. In the UK the incidence of aHUS is 0.41/per million/y [1] and the prevalence approximately 4.9 people per million population in Europe [2]. As there is the potential for patients with aHUS to relapse, lifelong treatment is recommended [3]. Until 2011, when eculizumab (a monoclonal antibody) was approved in Europe and the USA [4], plasma therapy and dialysis were the recommended treatments for aHUS but had little impact on disease morbidity and mortality and patients’ quality of life was poor [5]. Eculizumab is an effective treatment and in a genotype matched cohort improved 5-year end stage kidney disease free survival from 40% to 86% [1]. As eculizumab inhibits the body’s defence against certain infections there is a potential risk of a severe meningococcal infection [6–8]. The drug is also costly at £360,000 per year for an adult in the UK the cost, based on British National Formulary data. Eculizumab is administered by intravenous injection every two or three weeks. For people with aHUS managed with eculizumab there is the potential disruption of the onset of the illness, treatment, side effects, expectation of life-long treatment, and prophylactics.

Evidence is emerging that patients can stop eculizumab [9–11] and if they relapse treatment can be restarted with no evidence of long-term harm [12]. Patient and caregiver views on medication withdrawal have been explored in juvenile idiopathic arthritis [13] and rheumatoid arthritis [14] but little is known about those of aHUS patients on eculizumab withdrawal.

As part of a UK trial to establish whether a safety monitoring protocol could be an alternative management strategy to long-term treatment with eculizumab [15] we conducted an embedded qualitative study to explore patients’ or for children their parents’ views of treatment, withdrawing from treatment and the acceptable level of monitoring whilst off the drug. The findings will help to guide clinicians and patients/parents in their discussions in clinical consultations about eculizumab withdrawal.

## 2 Methods

This was an embedded qualitative study employing in-depth semi-structured interviews [16]. Due to the exploratory nature of the research a qualitative approach was considered appropriate [17]. As the aHUS population are geographically distributed across the UK, telephone interviews were more feasible and appropriate [18]; our PPI group agreed this approach. A topic guide was developed with the public contributor who was part of the trial team and research grant co-applicant.

### 2.1 Setting and recruitment

Adults and children with aHUS on eculizumab were identified from the National aHUS database [19]. Their medical records were assessed by the Newcastle clinical team and responsible local clinician to determine suitability for withdrawal [15]. Site clinical teams discussed the trial with eligible patients/parents when they attended for treatment. All were asked to consider participating in the qualitative study and given information and reply sheet to complete and return if they wished to discuss this with the researcher. The researcher followed this up with a telephone call/email to answer any questions and schedule an interview if they wished to proceed.

### 2.2 Data collection

Telephone interviews were conducted (by JL) at a time convenient to the participant. Interviews were digitally recorded and guided by a topic guide which this was used flexibly to enable participants to speak freely. Verbal consent was obtained at interview and recorded digitally and on paper. At the end of the interview withdrawal group participants were asked for permission to be approached again at the end of the two-year trial period for a second interview.

### 2.3 Analysis

There was no a priori theory, and we took an inductive approach to data collection and analysis [20]. Data were analysed thematically using a constant comparative method [21]. A framework was developed from a small number of transcripts, tested and amended accordingly, and the full data set then coded. A second level analysis involved a process of comparing and contrasting themes within and across interviews, and between the first and follow-up interviews. NVivo (Version 14) was used as a data management tool. Throughout the study findings were shared with the wider team, including lay members with direct experience of aHUS, for comment.

## 3 Findings

Between January 2019 and June 2022 eight adults (four withdrawals, four non-withdrawals) (Table 1) and 12 parents of children (Table 2) with aHUS (10 withdrawals, and two non-withdrawals) were interviewed from 9/15 participating hospital sites. Follow-up interviews were conducted with 11 of the 14 withdrawal group adults/parents.

**Table 1.** Adult’s age, sex, potential cause of onset and time since starting eculizumab.

| Withdrew from Eculizumab | Sex | Age (yrs) | Self-reported potential cause of onset | Time since starting eculizumab (years) |
| --- | --- | --- | --- | --- |
| Pat 01 | Female | 26-30 | Pregnancy | 3 |
| Pat 05 | Male | 21-25 | Gallstones | 2 |
| Pat 10 | Female | unknown | Pregnancy | 2.5 |
| Pat 21 | Female | 26-30 | Influenza | 3 |
| Did not withdraw from Eculizumab |  |  |  |  |
| Pat 02 | Female | 21-25 | Limb injury | 4 |
| Pat 03 | Female | 71-75 | Unsure | 8 |
| Pat 06 | Female | 26-30 | Virus | 5 |
| Pat 07 | Male | 55-60 | Swollen limb | 5 |

**Table 2.**
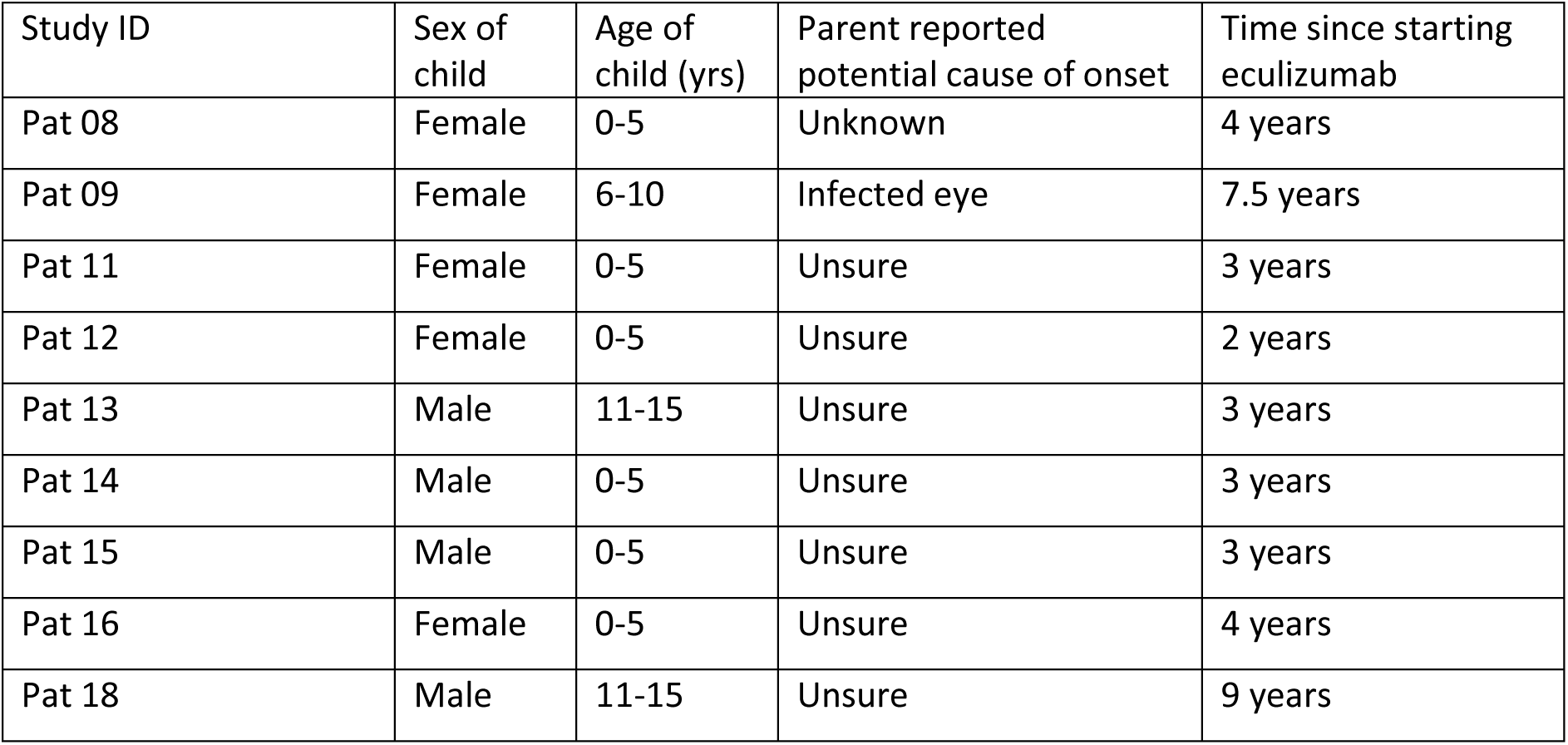
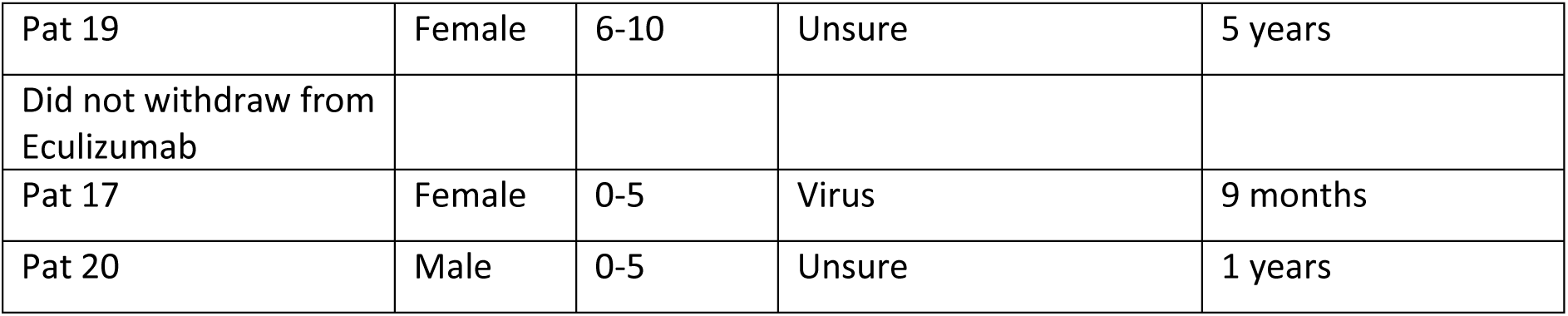
Children’s age, sex, potential cause of onset and time since starting eculizumab.

| Study ID | Sex of child | Age of child (yrs) | Parent reported potential cause of onset | Time since starting eculizumab |
| --- | --- | --- | --- | --- |
| Pat 08 | Female | 0-5 | Unknown | 4 years |
| Pat 09 | Female | 6-10 | Infected eye | 7.5 years |
| Pat 11 | Female | 0-5 | Unsure | 3 years |
| Pat 12 | Female | 0-5 | Unsure | 2 years |
| Pat 13 | Male | 11-15 | Unsure | 3 years |
| Pat 14 | Male | 0-5 | Unsure | 3 years |
| Pat 15 | Male | 0-5 | Unsure | 3 years |
| Pat 16 | Female | 0-5 | Unsure | 4 years |
| Pat 18 | Male | 11-15 | Unsure | 9 years |
| Pat 19 | Female | 6-10 | Unsure | 5 years |
| Did not withdraw from Eculizumab |  |  |  |  |
| Pat 17 | Female | 0-5 | Virus | 9 months |
| Pat 20 | Male | 0-5 | Unsure | 1 years |

Three key themes were elicited from the first stage interview data: trauma of illness onset; adjusting to lifelong eculizumab; and reasons for eculizumab withdrawal/non-withdrawal. The quotations are labelled ‘Ch’ Child, ‘Ad’ adult, ‘W’ Withdrawn and ‘NW’ Not withdrawn from eculizumab.

### 3.1 Theme 1: The onset of symptoms and receiving a diagnosis - ‘I was so ill. I thought I was on my way out’

Many participants described the onset of aHUS as traumatic and frightening. Some became emotional when giving their account. At onset, most adults experienced severe and sustained vomiting; for others it was sudden severe stomach pain, darkened urine, localised limb swelling or extensive bruising from a minor injury. In babies/children the symptoms were more varied though a common feature was extreme lethargy and sleepiness. Due to the rarity of aHUS other potential causes such as pre-eclampsia and meningitis were investigated. Adults and children underwent extensive tests, including lumbar puncture and kidney biopsy, and received treatments such as dialysis, blood transfusion and plasma exchange. A few were transferred to different hospitals, over weeks or even months. Several were diagnosed with aHUS within 24 hours and for others it ranged from one to four weeks, the longest was three months.

Some expressed fear of another episode ‘I think with this condition you are always anxious’ (P18-Ch-W). Uncertainties about the cause meant parents were vigilant if their child had any minor illness. Participants had become expert in, and spoke with confidence about, the routine test results and when there was cause for concern. Being aware of, and able to monitor, any changes gave a sense of control over the situation.

### 3.2 Theme 2: Adjusting to a lifetime on eculizumab - ‘We then started on the journey of regular infusions’

Participants were relieved to be offered eculizumab. After hospital discharge eculizumab was administered by infusion (ranging from 30 to 80 minutes) every two weeks and for a smaller number every three weeks. Some commented that the two weeks came around very quickly: ‘You felt like you were there every week’ (P15–Ch–W). Half had progressed to a nurse administering the drug at home, which saved a trip to hospital. However, this was not without its problems and on occasions had to be abandoned.

With babies/children there were issues initially with the cannula which, if their child became upset, was also stressful for parents. This became less of a problem over time but for some remained difficult for many years. Some adults and children were given more permanent vascular access (Portacath or peripherally inserted central catheter) to administer eculizumab and take blood without using needles. Two had experienced problems with semi-permanent access; one found it restricted bathing and leisure activities and another the central line had become infected.

#### 3.2.1 ‘Every time I wake up it takes a couple of hours to get going’ - the physical impact of eculizumab

Adults attributed a range of physical symptoms to the eculizumab. Tiredness was most cited; for several this was constant and for others lasted one or two days or passed if they were able to sleep post-infusion. With the fortnightly regime one person said they were *not tired* in only two weeks of each month. Other symptoms included leg and back pain, and headaches:

> *‘After eculizumab for two to three days it makes me feel very tired and I have body pain, leg pain and back pain. The leg pain is the worst. At night I even couldn’t move my legs in bed.’ **P01–Ad-W***

In very young children, lethargy, and tiredness post treatment was the main issue. Headaches, dizziness, aches and pains and slight gastrointestinal upset were also mentioned, which resolved relatively quickly.

A few experienced not too troublesome or no side effects. One person reported that after treatment ‘I literally get up and go home’ (P07–Ad-NW). For another, tiredness came on a few days *preceding* the eculizumab infusion, which they considered a sign they needed the drug:

> *‘You can almost tell though. I know when I need my treatment. Like I feel tired and more drained, more lethargic. I get sore legs so I know when it’s treatment time.’ **P06-Ad-NW***

#### 3.2.2 ‘We work around things’ – the social impact of taking eculizumab

The infusions caused general disruption to school, work and holiday. Those who experienced post-treatment tiredness felt they led a normal life only one week in two. The treatment, and any post-infusion tiredness, meant time off work and/or school. A few were self-employed and could more flexibly manage their time though this meant income loss . There were concerns they may be less desirable to future employers and about the stigma of providing proof for time off work.

Certain participants normalised treatment impact. One described taking an afternoon off work as a ‘pain’ but ultimately ‘no price to pay’ (P02-Ad-NW) and another that ‘it’s just one of those things’ (P17-Ch-W).

Taking longer holidays could be problematic though a few said with sufficient lead in time the hospital staff were able to support two-week holidays by gradually shifting treatment days to accommodate specific travel dates. However, some parents were fearful of being farther away from their care team should their child become ill.

In the non-withdrawal group holidays were less of an issue. They rarely went away on holiday, which was a preference rather than something imposed by their treatment.

> *‘We don’t have many holidays and I don’t let anything interfere with my treatment every fortnight. They can do it abroad if you want to but I see it a bit pointless. This drug is more important than a holiday.’ **P03-Ad–NW***

### 3.3 Theme 3 – Reasons for withdrawing/not withdrawing from eculizumab

#### 3.3.1 Withdrawing from eculizumab

Parents made the decision for their child to withdraw from eculizumab though two slightly older children were part of the discussion. Two adults disclosed that their families disagreed with their decision to withdraw. Most gave more than one reason for wishing to withdraw. The cost of the drug was a concern for many, but other reasons related to wanting a ‘normal’ life and issues with eculizumab.

##### ‘I just wanted to stop it so I can feel like I am a normal person’ – the wish to lead a normal life

Several participants talked of wanting a normal life for themselves or their child. What constituted a normal life varied between participants and was multifactorial. These factors included being able to travel for longer periods and further afield (an issue for those who had close family abroad), a life without regular medication and treatment side effects, and where their daily life and leisure time were not impacted.

One parent stated their child was ‘fed up with trips to the hospital.’ (*P13**-**Ch-W).* Parents had considered their child’s future, some as young as two years old, with lifelong medication. They wanted the chance to explore whether their child could lead a normal life without it. The opportunity to withdraw in the safety of a monitored trial was described as a ‘no lose’ situation with the guarantee of immediate eculizumab (that they know works) reinstatement should their child relapse. Finally. that regular test results, e.g. kidney function tests, had been normal for a long time reassured participants that now was the optimum time to withdraw from eculizumab.

> *‘All signs are good – I want to feel normal.’ **P01–Ad–W***

##### Issues with eculizumab

###### Concerns about long-term eculizumab

Uncertainty about the long-term effects of eculizumab and the requirement for lifelong treatment was a cause of concern. There were also fears about eculizumab being an immunosuppressant and the higher risk of meningitis and pneumonia. This was a major concern for one parent for the four years their child had been on eculizumab and a reason for withdrawing. They felt with the monitoring protocol detecting an aHUS relapse would be easier than detecting that their child had meningitis.

###### Is the treatment warranted? ‘If I don’t have to be on it, I don’t want to be on it’

Participants wanted to know whether eculizumab was needed, particularly long-term. This was driven to a large degree by the clinical team’s uncertainty about the aHUS diagnosis. Diagnostic uncertainty as a driver for withdrawal was coupled with a dislike of long-term medication where it is not fully justified, and its impact on their bodies, particularly young children. One parent thought even a short break from eculizumab would be beneficial as they could stop prophylactics. Although mentioned by only one parent, there was uncertainty over the optimum eculizumab dose, and because of this withdrawal was suggested as a viable option. When their kidney function and other related tests had been stable for a long time adults questioned whether they needed eculizumab.

One participant disliked needles and described the fortnightly cannula insertion as ‘traumatising’. Their decision to withdraw was driven primarily by eculizumab side effects.

> *‘My hair was thinning and I was getting migraines and all these things so I heard about this study where you could come off it and I was like, ‘Yes please!’. **P21–Ad- W***

#### 3.3.2 Reasons for not withdrawing from eculizumab

The six non-withdrawal participants all stated that having the opportunity to do so safely was a good idea. Of these, two had hoped to withdraw (reasons included in the previous section) but the clinical team advised against it. The first had another condition that caused proteinuria and for the second, a child, there were concerns about the impact of the regular blood tests on their veins. For these reasons the clinical team decided monitoring disease activity whilst off eculizumab would be difficult.

For the remaining four participants the reasons for not withdrawing varied. The one parent commented it was a relatively short period since their child had commenced eculizumab and they had had complications following surgery for an unrelated condition. Despite the proposed monitoring plan, they felt detecting a relapse would be difficult as their child was too young to properly communicate they were unwell.

The three adult participants were concerned about relapse and being as ill as they were initially. All had adapted into living with eculizumab and did not want this disrupted. The first assumed withdrawal would lead to kidney failure, time off work and income loss; the three weeks spent in hospital were very difficult. For the second, being on eculizumab gave them peace of mind; if they withdrew, they believed with every minor health issue experienced they would be terrified they were relapsing. With debilitating co-morbid conditions and being much older they felt they may not survive a relapse. The third as a constant ‘worrier’, was anxious about the uncertainty of *when* relapse might occur and did not want to live with the constant fear every time they had a cold that the aHUS had returned. They also disliked the idea of having their portacath removed and reinserted elsewhere if treatment was resumed. Lastly putting themselves at risk of a relapse and being ill would negatively affect their family.

> *‘Also I don’t want to be like, “Okay I‘m going to do this” …because it doesn’t just affect me, it affects all my family. That’s the hardest thing of being ill. At the time when I relapsed it wasn’t me it was watching them, watching their reactions. It’s hard to discuss.’ **P06-Ad-NW***

A final concern was whether in the two-year period there may be a decision to no longer fund the drug. In that eventuality, they believed participants could be left without an effective treatment.

The adult non-withdrawals were interested in the results of the main trial and would consider eculizumab withdrawal if there was ‘definite’ evidence of its safety. However, this evidence would have to demonstrate that someone with the same genetic mutations had safely withdrawn, ‘because of all of the factors in play’ (P06–Ad-NW).

### 3.4 Life after eculizumab

Participants interviewed after two years had not restarted eculizumab. The trial monitoring protocol had given reassurance and a sense of control over what was happening. Only one adult thought the initial frequent monitoring was excessive. Most participants were happy that the number of tests/hospital visits had reduced over time. One parent missed the support from the research nurses during monitoring visits, particularly discussing their anxieties about their child.

Two overarching themes were elicited from the data, ‘Returning to normality?’ and ‘A new normal’.

#### 3.4.1 Return to normality?

Eculizumab withdrawal had positively impacted on participants’ lives in three main ways. The first related to the side effects attributed to eculizumab. One participant said these had disappeared almost overnight. For another it was taking time to return to their pre aHUS state, but the extreme fatigue was ‘improving day by day’. The second was the changes to their lives of no longer having treatment. Though taking the time out of work and/or school had become part of their routine, it was liberating to no longer have to do so.

> *‘We’ve just booked to go away because then [child] is going to finish the trial so we thought we’d do a whole celebratory thing for her to kind of say she’s free now.**’ P11–Ch-W***

One parent said after stopping prophylactic penicillin their child now recovers more quickly from minor illnesses. It was also a relief to no longer have the psychological impact of witnessing their child’s distress during treatment, for example cannulation:

> *‘From a sort of mental perspective it’s done us all a world of good’. **P15-Ch-W***.

Third, there were many references equating the cessation of treatment to being ‘normal’. This was due to less disruption to their daily lives. Also, taking the drug had been a reminder they had an illness and, particularly for children, were different to others.

#### 3.4.2 A new normal

Despite the positive impact of withdrawal, one participant said coming off treatment had triggered a rollercoaster of emotions. Concerns about a relapse whilst off eculizumab continued:

> *‘Now I’m off the drug I feel like a ticking time bomb, it could happen again at any time’. **P21–Ad–W***

Early in the trial parents admitted to closely observing their child and continually looking for signs of a relapse. Most said this had lessened over the two-year period, but their anxiety remained in the background, and a few were constantly worried. One parent had noticed a change in their child’s behaviour, being more withdrawn and in a world of their own. They were unsure whether this was related to treatment withdrawal or changes as they enter adolescence.

Participants had a sense of control and greater resolve that they/their child could confidently withdraw from the treatment, and were assured they could deal with a relapse. One participant said they took greater care of themselves to avoid becoming ill and triggering a relapse. Regular testing (bloods), having a hospital contact, knowing the procedure should they/their child relapse and that eculizumab can immediately be re-started was of major importance.

Participants acknowledged long-term follow-up was required and parents thought the current monthly monitoring might be reduced over time. Some said they would be happy if this were once or twice a year, providing they had an immediate route into hospital when needed. The majority were waiting to discuss a care plan with their local consultant, but one participant had agreed theirs:

> *‘Every six months and that’s for my lifetime. …, it’s not a bother and it’s actually quite nice knowing I am being monitored and I’m not just being left.’ **P21–Ad-W***

## 4 Discussion

In this qualitative study we explored adult aHUS patients’ and for children, their parents’ experiences of the onset of aHUS, of eculizumab and its withdrawal. Little has been reported from the perspectives of adults and (parents of) children with aHUS.

### 4.1 Decisions to withdraw from, or remain on, eculizumab

Key motivations for withdrawal participants were to attain some normalcy in their lives and concerns about potentially unwarranted long-term medication. Side effects were an issue particularly for adults, which in other conditions impact on patients’ daily lives [22] and are a key determinant in patients’ health related quality of life [23]. Adults were concerned about their status as a valued workforce member and the stigma of being considered a malingerer because of work absences for treatment. Parents worried about their child being different to their peers and the questions they faced - or would face in the future – when they took time off school for treatment. Withdrawing from eculizumab was considered an opportunity to be a ‘normal’ person. Similar findings on reasons for withdrawal/non-withdrawal of medication have been reported in studies conducted with patients/caregivers with juvenile idiopathic arthritis [13] and rheumatoid arthritis [14]. With aHUS, monitoring and being able to quickly reinstate eculizumab - and knowing it is effective - should they or their child relapse provided reassurance.

The impact of the onset of aHUS was a key driver for *declining* eculizumab withdrawal. It had resulted in a changed existence of rarely being ill and no experience of hospital, to living with the ‘ticking time bomb’ threat of a potential relapse, constantly watching for anything they feel may act as a trigger and monitoring any changes in their regular tests. Their thoughts about eculizumab withdrawal focused heavily on a relapse and the subsequent disruption. Adults in this group normalised and reframed disruption to work and holidays of the eculizumab regime.

### 4.2 Two years post-withdrawal

After two years withdrawal from treatment adults and parents had regained some degree of normality, almost to what they were previously. Parents talked positively of the difference it had made to their child and family of not having the treatment and attending hospital on a fortnightly basis. Adults no longer experienced the debilitating side effects of the treatment, or these were diminishing over time, which meant they could function normally.

All acknowledged the need to be monitored by the local clinical team for the foreseeable future. Adults/parents had a greater sense of control knowing what to expect and do should they or their child relapse. Parents were cognisant that their child, as they grow older, will have to understand the condition and be aware of the potential triggers of a relapse. This all constituted a new normal.

### 4.3 Limitations

One limitation is the low number of non-withdrawal qualitative study participants (6/11). Second, two of the four withdrawal adults were lost to follow-up. Although the data from the two-year withdrawal period does reflect the views of adult patients, the majority were parents of children. However, as children/parents face a longer time on eculizumab than adults this is an important group to capture.

## 5 Conclusions

Most experienced a frightening ordeal at the onset and were grateful that eculizumab had been prescribed. However, over time the frequency, route of administration and side effects of eculizumab had become problematic for some. Discussions about eculizumab withdrawal should be informed by an understanding of their experiences of aHUS onset. Withdrawal generated anxiety, and support to alleviate fears in the early stages would be beneficial. Evidence from the main trial on the optimum monitoring, successful withdrawal, and recovery time where eculizumab was reinstated may provide reassurance to those who are uncertain.

## Data Availability

All data produced in the present study are available upon reasonable request to the authors

## Acknowledgements

We would like to thank the staff at the nine participating hospital sites for their help recruiting to the qualitative study. We are grateful to the adults with aHUS and the parents of children with aHUS for their valuable contributions to this study, for taking the time to participate in the initial and follow-up interviews and telling their stories.

## Statements and declarations

### Ethics approval

A favourable ethical opinion was obtained from North East-Tyne & Wear South Research Ethics (18/NE/0113)

### Availability of data and material

It is not possible for the data from this study to be made available.

### Authors’ Contributions

JL and NS devised the study design and the topic of exploration for the qualitative component. JL conducted the interviews, analysed the data and drafted the manuscript. NS reviewed and contributed to the final manuscript.

---

**Figure 1.**
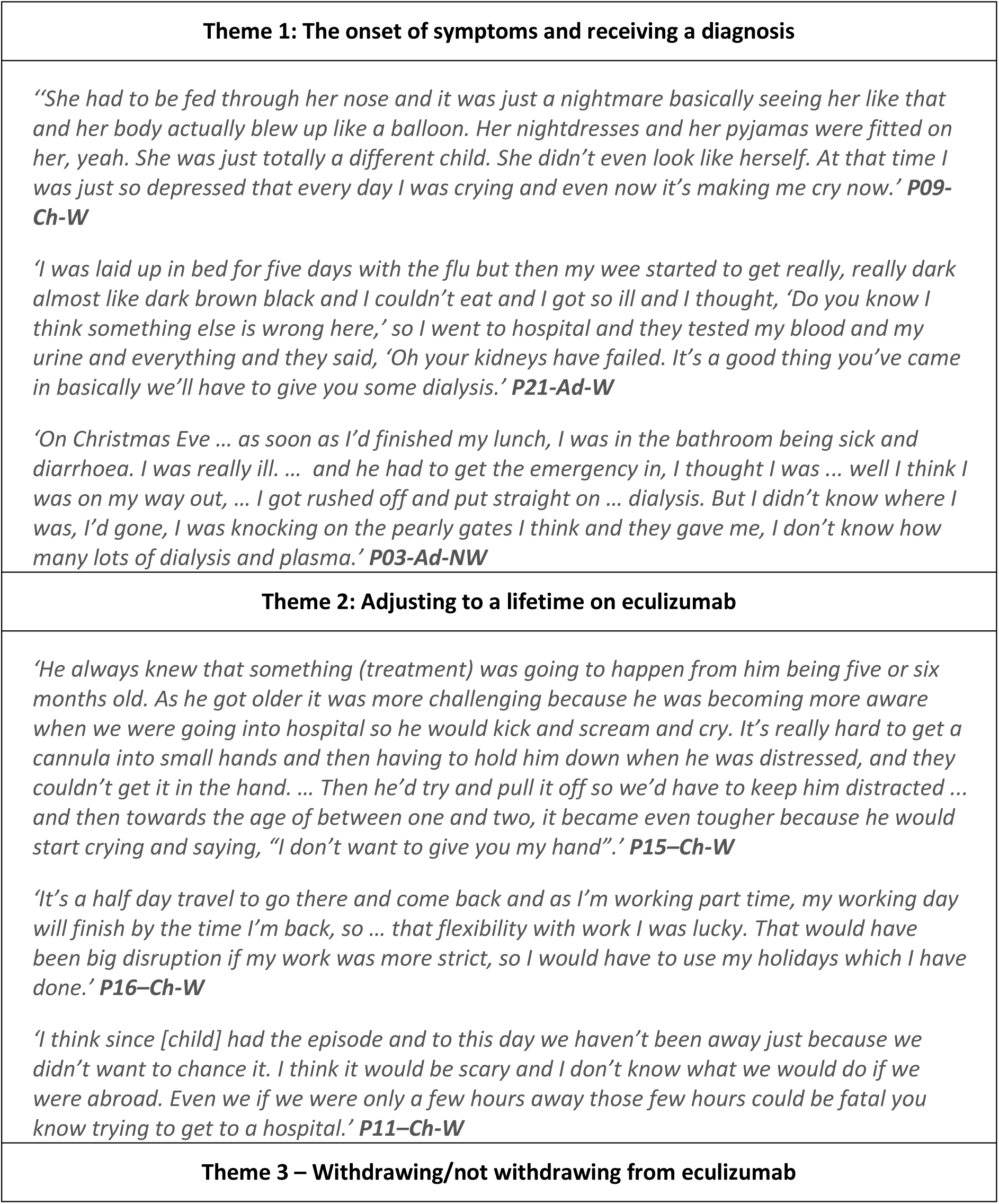
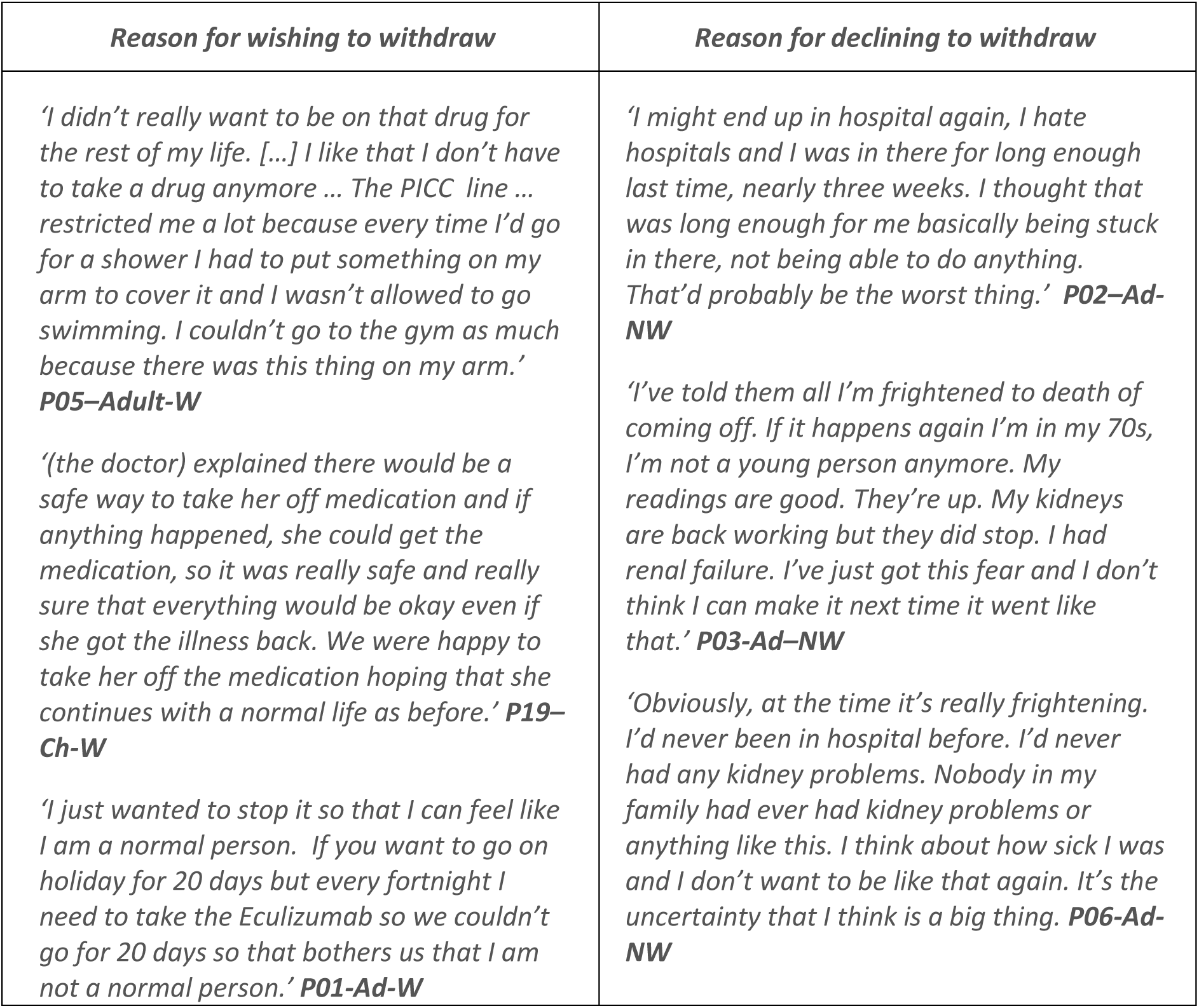
Themes 1-3: Participant quotations

**Figure 2.**
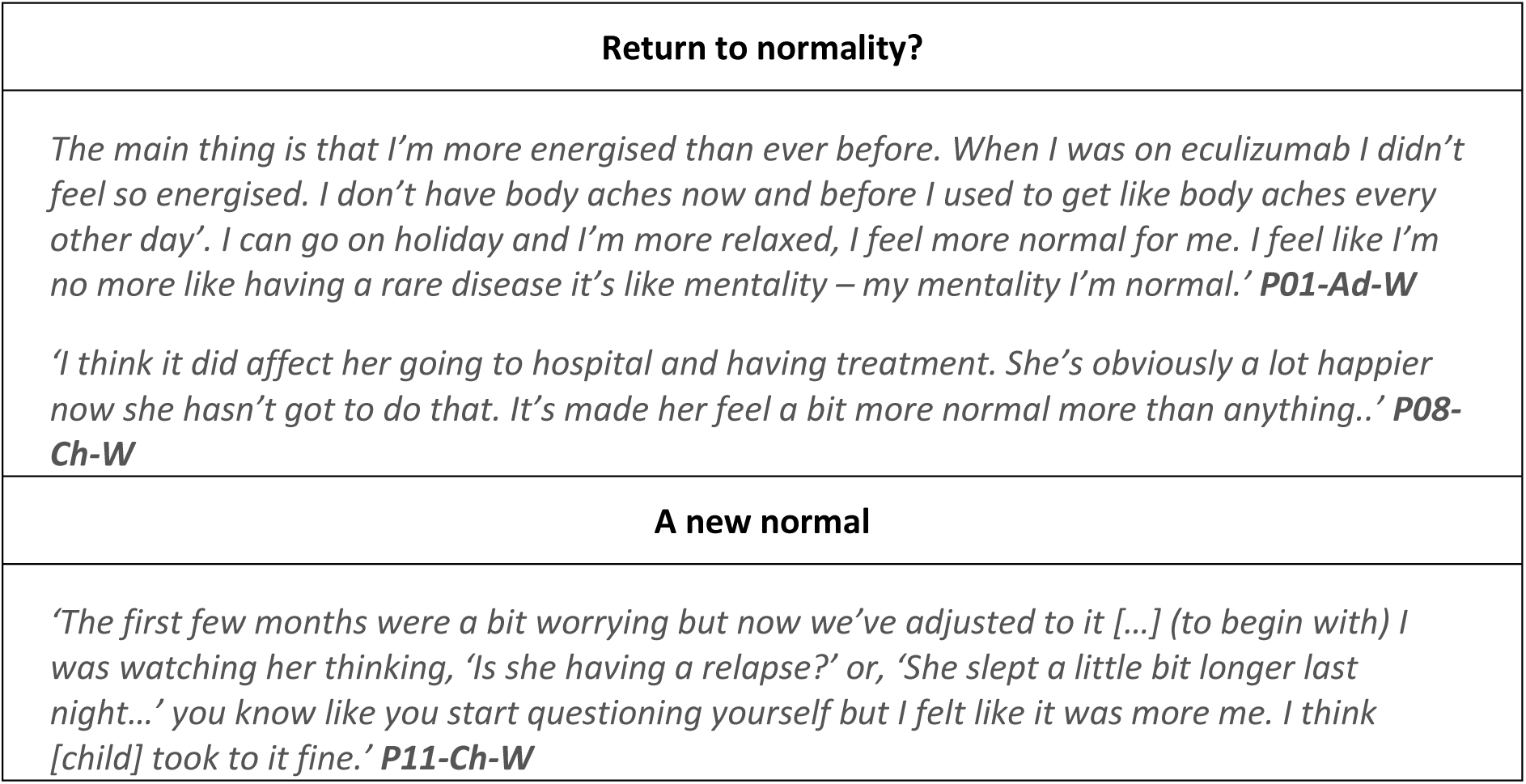
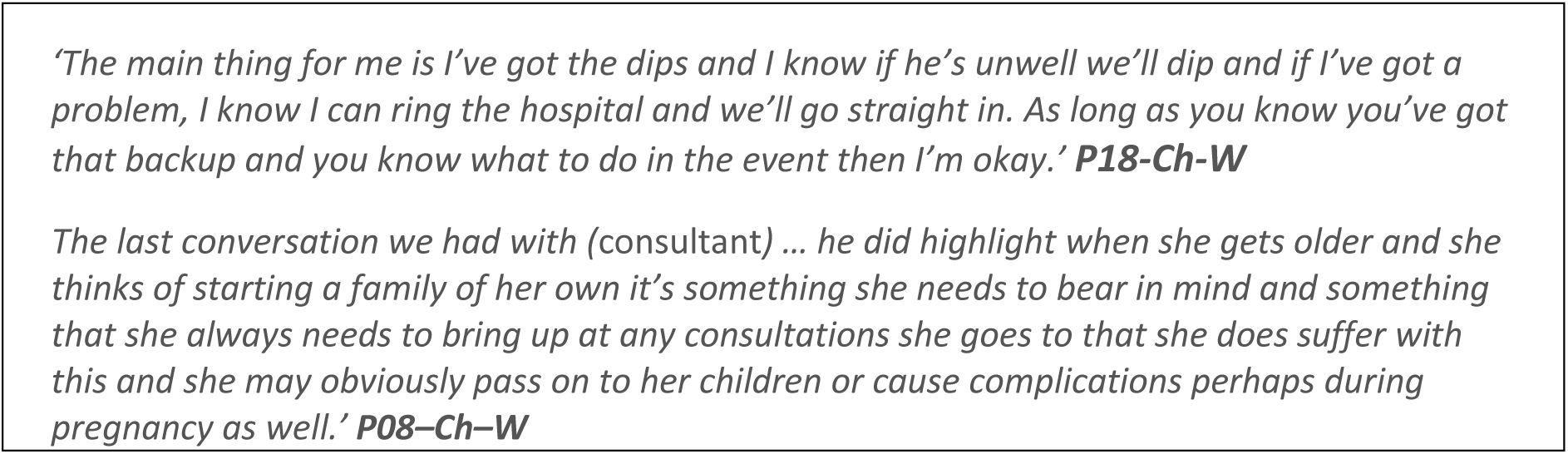
Life after eculizumab: participant quotations

